# The impact of multimodal cognitive rehabilitation on executive functions in older adults with traumatic brain injury (TBI)

**DOI:** 10.1101/2020.10.26.20220012

**Authors:** Eduardo Cisneros, Véronique Beauséjour, Elaine de Guise, Sylvie Belleville, Michelle McKerral

**Affiliations:** Centre for Interdisciplinary Research in Rehabilitation of Greater Montreal (CRIR)-IURDPM, CIUSSS du Centre-Sud-de-l’Île-de-Montréal, Montreal, QC, Canada; Department of Psychology, Université de Montréal, Montreal, QC, Canada; Research Institute of the Montreal University Hospital Centre, Montreal, QC, Canada; Research Centre of the Institut universitaire de gériatrie de Montréal, Montreal, QC, Canada

**Keywords:** traumatic brain injury, aging, cognitive rehabilitation, executive functions, self-awareness

## Abstract

**Objectives:** This study evaluated the impact of a multimodal cognitive rehabilitation intervention, the Cognitive Enrichment Program (CEP), on executive functioning (EF) and resumption of daily activities following traumatic brain injury (TBI) in older individuals, in comparison to an active control group having received holistic rehabilitation as usual care.

**Methods:** The CEP’sexecutive function module included planning, problem solving, and goal management training, as well as strategies focusing on self-awareness. Effectiveness was evaluated by psychometric tests (Modified Six Elements Task-adapted – MSET-A, D-KEFS *Sorting test* and *Stroop four-color version*), while generalization was measured through self-reported questionnaires about daily functioning (Dysexecutive Functioning Questionnaire – DEX, *Forsaken daily life activities*). Measures were obtained before and after intervention, and six months later.

**Results:** ANCOVA results showed significant group-by-time interactions on *Tackling the 6 subtasks* and *Avoiding rule-breaking* measures of the MSET-A, with moderate effect sizes. Despite improvements in Sorting and Stroop scores, there were no group-by-time interaction on these measures. DEX generalization measure showed a significant reduction in patient/significant other difference on the *Executive Cognition* subscale. There was a reduction in the number of *Forsaken daily life activities* in the experimental group compared to controls which was not significant immediately after CEP, but that was significant six months later.

**Conclusions:** Our study shows that older adults with TBI can improve their executive functioning with a positive impact on everyday activities after receiving multimodal cognitive training compared to an active control group.

## Introduction

Cognitive rehabilitation programs for older individuals having sustained a traumatic brain injury (TBI) are lacking [1,2], even if this population is growing and presents multiple vulnerabilities [3]. Normal aging is accompanied by neurophysiological, neuroanatomical and physical decline that can negatively influence neurocognitive functioning, including attention, memory, and executive functions (EF) [4,5]. These vulnerabilities can worsen clinical outcome when a TBI is sustained during older adulthood [3,5].

In spite of age-related brain function decline, the frequent presence of general age-related health comorbidities (diabetes, hypertension, hypercholesterolemia, etc.) and the collateral effects of medication on cognition or mood, some studies suggest that older individuals with TBI could benefit from intensive rehabilitation programs and positive impacts on their level of independence even if their length of stay in clinical settings is longer [6-8].

Among cognitive symptoms produced by TBI memory, attention and executive dysfunctions are probably the most disabling sequelae which in many cases disturb the individual’s ability to live independently [3,9,10]. Executive functions (EF) represent a multidimensional concept including initiating, planning, organizing, solving problems, choosing among different options, self-regulating one’s own behavior, forming concepts, maintaining goals until attaining them in a satisfactory way, as well as adjusting plans or goals to adapt to environmental and personal changes [11]. These higher-level functions include also self-awareness abilities, emotional and behavioral regulation and social adjustment [12-13]. By their anatomical position and their interconnectivity with other areas of the brain, frontal lobes, highly involved in EF organization, are more vulnerable to diffuse injuries as TBI [11,14,15]

In order to approach rehabilitation possibilities in older adults with TBI, it is important to learn from research conducted with healthy older individuals as well as with adults with TBI. Meta-analyses of cognitive rehabilitation in these populations show significant effectiveness on cognitive functioning, including EFs, and generalization of gains to real-world activities [16,17]. This is more likely to be attained if rehabilitation programs address multiple EFs rather than a unique or specific EF. However, low representation of older TBI population limit the scope of recommendations [18-20].

Stuss and colleagues [21] published a series of reports from a randomized trail conducted with normal ageing people showing non pathological cognitive complaints, which aimed to assess the effectiveness of a multidomain cognitive training program. This program was composed of three modules, one for memory functions, the second for EF training, and the last one for psychosocial dimensions. In the study reporting the specific effects on EF, Levine et al [22] used an adapted version of the GMT programThe effectiveness of training was measured through questionnaires, neuropsychological tests and by life-like tasks. Large effect sizes were obtained for global measures of EF and for two of the specific measures, namely, task strategy and monitoring/error correction. Effect size was moderated by task engagement. A significant effect was found for the Dysexecutive questionnaire (DEX). The effects were maintained 6 months after the end of the program.

Later, Levine et al. [23] conducted a controlled trail with a sample of 19 acquired brain-injured (ABI: stroke, TBI and tumors) younger adults using an expanded version of the GMT which was based on the theory of sustained attention [24]. According to this theory, activation of the frontal-parietal-thalamic attention system is required to endogenously sustain goals in working memory. Following ABI, those systems are affected and endogenous attention is perturbed, leaving individuals cue-dependent. The GMT main strategy thus consists of periodically stopping ongoing behavior to self-monitor performance and define and adjust goals. Their study, comparing GMT effectiveness in ABI to a control group showed significant and specific effects on attention, maintaining awareness and self-regulation [23]. Recently, meta-analyses [25-26] showed that GMT can enhance improvement in executive functioning in ABI patients.

Spikman and colleagues developed a multimodal program [27] that exclusively addressed executive dysfunction. This program included, among others, the Planning Method, the Problem-Solving Method and GMT. They assessed its effectiveness with ABI (stroke, TBI and brain tumors) in young and middle-aged adults through a multicenter randomized controlled trail [28]. Effectiveness of this 20-24 sessions rehabilitation program was assessed using life-like tasks, neuropsychological tests, observation and questionnaires. The experimental ABI group showed better abilities to set and accomplish realistic goals, planning, initiating, and regulating real-life tasks, and resuming previous roles with respect to work, social relationships, leisure activities, and mobility.

To our knowledge, there are no studies evaluating the effects of comprehensive rehabilitation programs on, among others, EFs in TBI older individuals. This paper reports on the EF component of a newly designed cognitive rehabilitation program, the Cognitive Enrichment Program (CEP), which was tailored for individuals who sustain a TBI in later adulthood. The CEP is a 12-week multimodal intervention structured into three modules designed to address cognitive problems resulting from TBI, as well as age-related cognitive issues in the following domains: self-awareness, attention and episodic memory, and EF. The EF module integrates training on planning, problem solving, goal management and self-regulation strategies.

We evaluated the effectiveness of the CEP for enhancing EF in older adults with TBI, as assessed with three neuropsychological EF measures as primary outcomes, as well as a self-report questionnaire of daily executive functioning and a measure of daily life activities as generalization measures of real-world EF performance. We hypothesized that targeted EF training included in the CEP would lead to improvements on psychometric EF measures as well as in self-reported executive cognition and resumption of daily activities in a trained group of older individuals with TBI, whereas this would not be the case for a comparable TBI group who did not receive the CEP intervention. A secondary objective was to assess stability of change six months after the end of training.

## Material and Methods

Results presented in this paper are part of a larger controlled clinical trial (study registered at ClinicalTrials.gov, Identifier: NCT04590911). Experimental design, participant selection and allocation, general CEP intervention framework, as well as detailed assessment procedures are described in Cisneros et al. and its accompanying Supplementary material [29] (this issue).

### Procedure

The CEP consists of three intervention modules, Introduction and self-awareness, Attention and memory (strategies detailed in Cisneros et al. [29]; this issue), and Executive functions (EF), which include a total of 24 sessions of 90 minutes twice weekly within a 12-week time frame [29]. The CEP module targeting EF consists of three approaches: Planning Method (PM), Problem-Solving Method (PSM), and GMT. The PM was derived from Spikman et al.’s program [27] that we translated to French from Dutch with the collaboration of the authors. Emphasis is put on organizing steps and anticipating of time, as well as human and material resources in order to resolve a complex situation. The PSM is an adaptation of von Cramon et al.’s [30] and Spikman et al.’s strategies [27,28]. Participants first learn to verbalize questions during problem-solving and then this process is progressively faded-out. They must select the significant information pertaining to the problem, formulate the problem, generate as many solutions as possible, select and execute the most appropriate solution to a problem, and verify its effectiveness. The GMT is mainly based on Spikman’s [27,28] and Levine’s [22,23] adaptations of the original GMT. Exercises for each method gradually increase in difficulty from easier-fictive exercises to more complex and real-life tasks.

In addition to the specific EF interventions, strategies targeting self-awareness were implemented during CEP intervention: general discussion about self-awareness after TBI, keeping a list of cognitive strengths-difficulties [27], feedback about progress on overcoming difficulties, and discussion about actual self-awareness difficulties using the *Reaction* tool. The latter is a card-based instrument developed at the TBI program of the Lucie-Bruneau Rehabilitation Centre and consists of a set of 24 real-life situations describing different examples of self-unawareness [31]. Participants are asked to identify, for each card, the awareness problem, and to share if they had lived a similar situation.

### Assessment times

Evaluations were conducted at three time-points: baseline (pre-intervention – T0); 14-weeks (post-intervention – T1); 6-months post-intervention (follow-up – T2).

### Primary outcome measures

We adapted Crépeau et al.’s [32] version of the original Six Elements Test [33] to a paper-pencil format, named the ***Six Elements Task-Adapted (SET-A)***. Five measures were used: ***Tackling the 6 subtasks, Inter-task balance, Avoiding rule-breaking, Checking time***, and ***Efficient behavior***. We also calculated the ***Total score*** of the five measures of the SET-A, as well as the ***Total number of points*** earned. See Supplementary material for more details on the measures used.

The ***Sorting Test*** from the Delis-Kaplan Executive Function System (D-KEFS) [35] was also used as a primary outcome measure. ***Confirmed Correct Sort Total (CCS)*** raw score, ***Free Sorting Description (FSD)*** total raw score, and ***Time-Per-Sort Ratio (TSR)*** were calculated.

The other primary outcome measure was the ***Stroop*** four-color version [36], where ***Inhibition*** and ***Flexibility*** conditions were used.

### Generalization measures

Self-awareness of executive difficulties was used as a measure of generalization by calculating the difference between participants’ and a significant other’s scores on the three subscales of the **Dysexecutive Questionnaire (DEX)** [37], as proposed by Simblett et al [38]: ***Executive cognition, Behavioural-Emotional Self-Regulation***, and ***Metacognition***.

Self-reported ***Forsaken Daily Activities*** from the Client’s Intervention Priorities tool [39] were thus used as a measure of generalization to everyday life.

### Control measure

The ***Similarities*** subtest from the WAIS-III was chosen as a control measure since it represented cognitive factors not related to EFs targeted by the CEP program. We did not expect any changes for this variable.

### Statistical analysis

The statistical approach used is described in Cisneros et al. [29] (this issue).

## Results

Demographic and clinical characteristics of the two groups can be found in Table 2 of Cisneros et al. [30] (this issue).

**Table 1.**
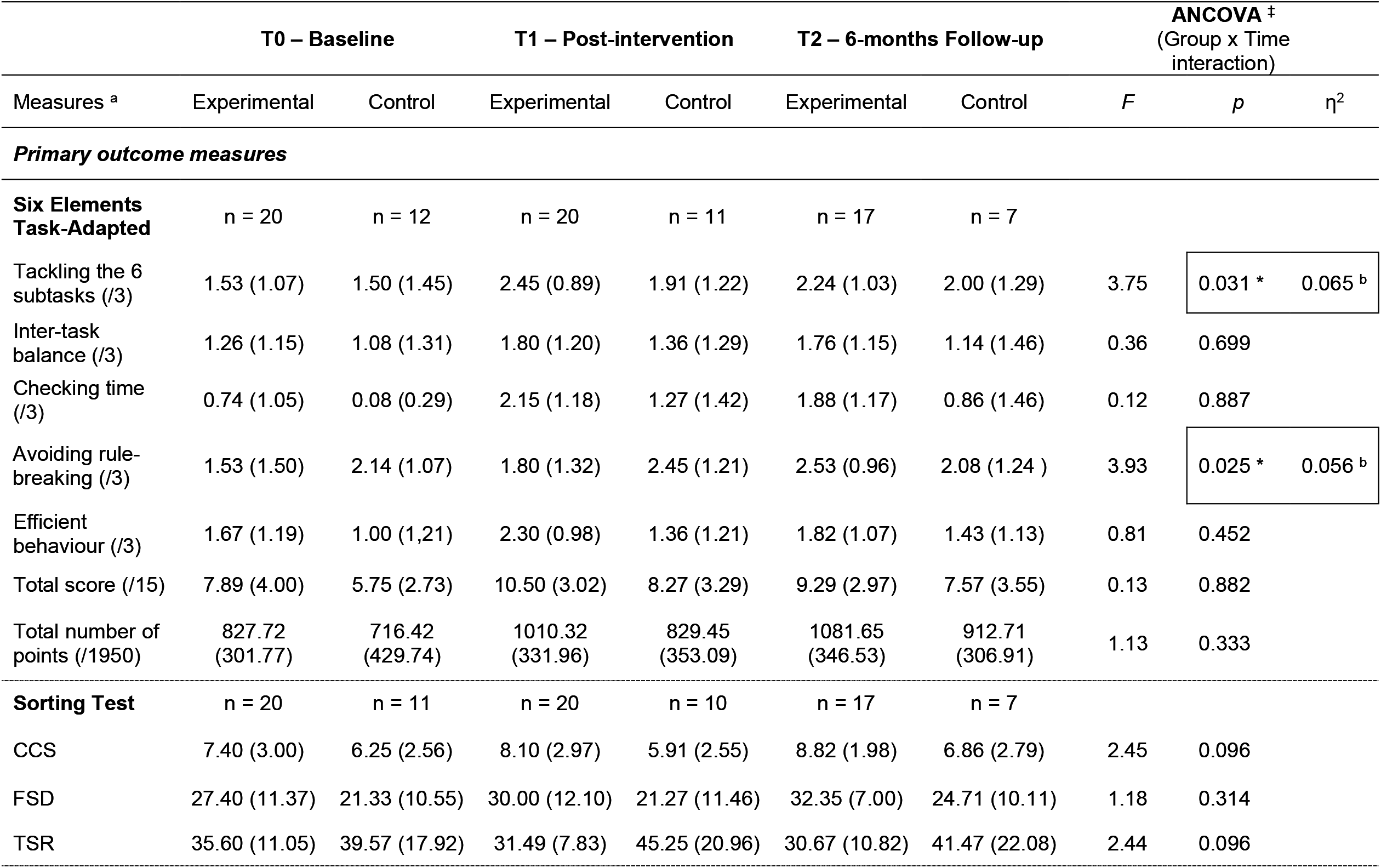
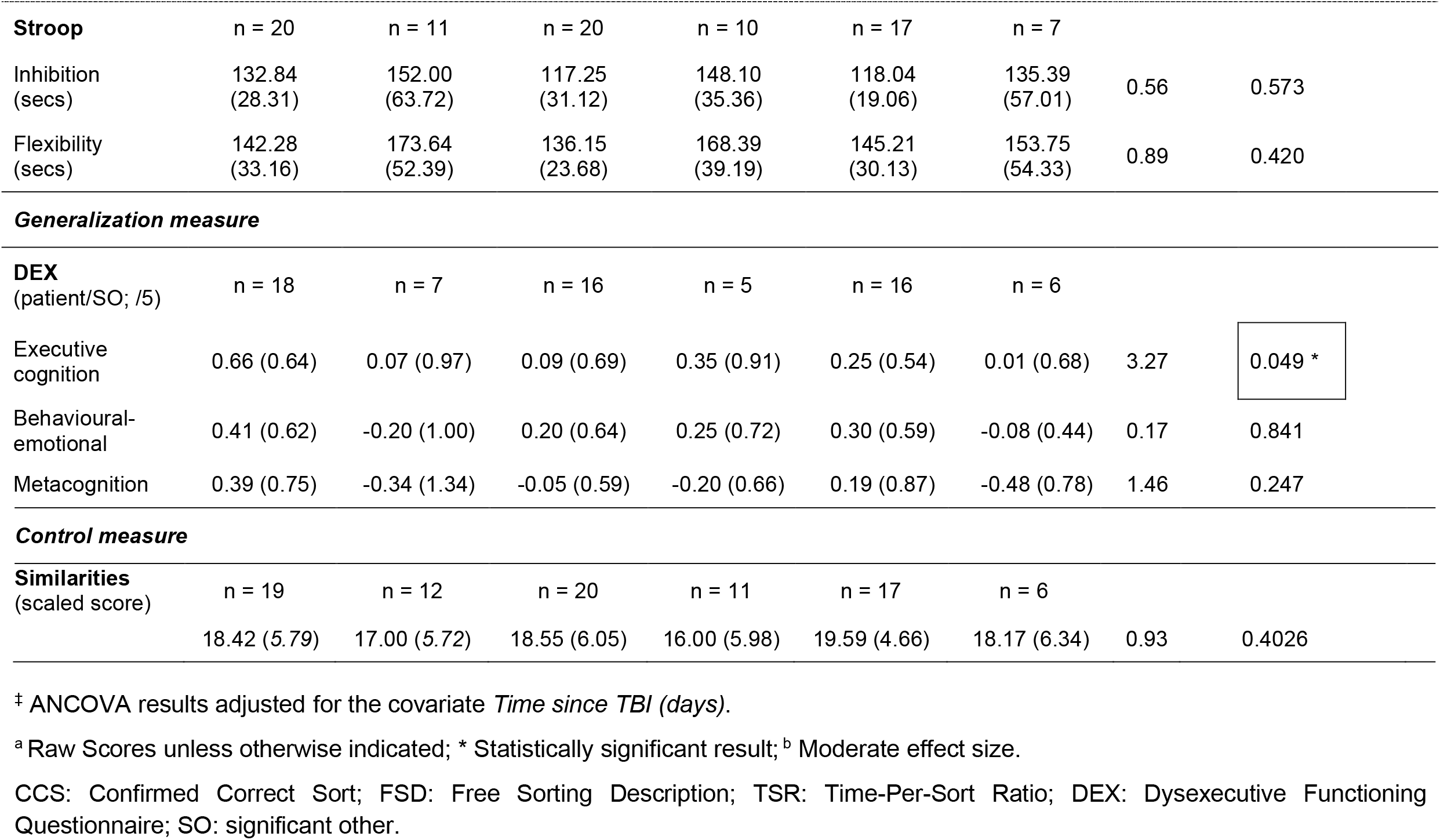
Means (SD) for the different measures at the three assessment times, and ANCOVA results.

| Measures <sup>a</sup> | T0 – Baseline |  | T1 – Post-intervention |  | T2 – 6-months Follow-up |  | ANCOVA <sup>‡</sup><br>(Group x Time interaction) |  |  |
| --- | --- | --- | --- | --- | --- | --- | --- | --- | --- |
|  | Experimental | Control | Experimental | Control | Experimental | Control | F | p | η<sup>2</sup> |
| **Primary outcome measures** |  |  |  |  |  |  |  |  |  |
| **Six Elements Task-Adapted** | n = 20 | n = 12 | n = 20 | n = 11 | n = 17 | n = 7 |  |  |  |
| Tackling the 6 subtasks (/3) | 1.53 (1.07) | 1.50 (1.45) | 2.45 (0.89) | 1.91 (1.22) | 2.24 (1.03) | 2.00 (1.29) | 3.75 | 0.031 * | 0.065 <sup>b</sup> |
| Inter-task balance (/3) | 1.26 (1.15) | 1.08 (1.31) | 1.80 (1.20) | 1.36 (1.29) | 1.76 (1.15) | 1.14 (1.46) | 0.36 | 0.699 |  |
| Checking time (/3) | 0.74 (1.05) | 0.08 (0.29) | 2.15 (1.18) | 1.27 (1.42) | 1.88 (1.17) | 0.86 (1.46) | 0.12 | 0.887 |  |
| Avoiding rule-breaking (/3) | 1.53 (1.50) | 2.14 (1.07) | 1.80 (1.32) | 2.45 (1.21) | 2.53 (0.96) | 2.08 (1.24 ) | 3.93 | 0.025 * | 0.056 <sup>b</sup> |
| Efficient behaviour (/3) | 1.67 (1.19) | 1.00 (1,21) | 2.30 (0.98) | 1.36 (1.21) | 1.82 (1.07) | 1.43 (1.13) | 0.81 | 0.452 |  |
| Total score (/15) | 7.89 (4.00) | 5.75 (2.73) | 10.50 (3.02) | 8.27 (3.29) | 9.29 (2.97) | 7.57 (3.55) | 0.13 | 0.882 |  |
| Total number of points (/1950) | 827.72 (301.77) | 716.42 (429.74) | 1010.32 (331.96) | 829.45 (353.09) | 1081.65 (346.53) | 912.71 (306.91) | 1.13 | 0.333 |  |
| **Sorting Test** | n = 20 | n = 11 | n = 20 | n = 10 | n = 17 | n = 7 |  |  |  |
| CCS | 7.40 (3.00) | 6.25 (2.56) | 8.10 (2.97) | 5.91 (2.55) | 8.82 (1.98) | 6.86 (2.79) | 2.45 | 0.096 |  |
| FSD | 27.40 (11.37) | 21.33 (10.55) | 30.00 (12.10) | 21.27 (11.46) | 32.35 (7.00) | 24.71 (10.11) | 1.18 | 0.314 |  |
| TSR | 35.60 (11.05) | 39.57 (17.92) | 31.49 (7.83) | 45.25 (20.96) | 30.67 (10.82) | 41.47 (22.08) | 2.44 | 0.096 |  |

|  |  |  |  |  |  |  |  |  |
| --- | --- | --- | --- | --- | --- | --- | --- | --- |
| **Stroop** | n = 20 | n = 11 | n = 20 | n = 10 | n = 17 | n = 7 |  |  |
| Inhibition (secs) | 132.84 (28.31) | 152.00 (63.72) | 117.25 (31.12) | 148.10 (35.36) | 118.04 (19.06) | 135.39 (57.01) | 0.56 | 0.573 |
| Flexibility (secs) | 142.28 (33.16) | 173.64 (52.39) | 136.15 (23.68) | 168.39 (39.19) | 145.21 (30.13) | 153.75 (54.33) | 0.89 | 0.420 |
| **Generalization measure** |  |  |  |  |  |  |  |  |
| **DEX**<br>(patient/SO; /5) | n = 18 | n = 7 | n = 16 | n = 5 | n = 16 | n = 6 |  |  |
| Executive cognition | 0.66 (0.64) | 0.07 (0.97) | 0.09 (0.69) | 0.35 (0.91) | 0.25 (0.54) | 0.01 (0.68) | 3.27 | 0.049 * |
| Behavioural-emotional | 0.41 (0.62) | -0.20 (1.00) | 0.20 (0.64) | 0.25 (0.72) | 0.30 (0.59) | -0.08 (0.44) | 0.17 | 0.841 |
| Metacognition | 0.39 (0.75) | -0.34 (1.34) | -0.05 (0.59) | -0.20 (0.66) | 0.19 (0.87) | -0.48 (0.78) | 1.46 | 0.247 |
| **Control measure** |  |  |  |  |  |  |  |  |
| **Similarities**<br>(scaled score) | n = 19 | n = 12 | n = 20 | n = 11 | n = 17 | n = 6 |  |  |
|  | 18.42 (5.79) | 17.00 (5.72) | 18.55 (6.05) | 16.00 (5.98) | 19.59 (4.66) | 18.17 (6.34) | 0.93 | 0.4026 |

‡ ANCOVA results adjusted for the covariate *Time since TBI (days)*.
<sup>a</sup> Raw Scores unless otherwise indicated; \* Statistically significant result; <sup>b</sup> Moderate effect size.
CCS: Confirmed Correct Sort; FSD: Free Sorting Description; TSR: Time-Per-Sort Ratio; DEX: Dysexecutive Functioning Questionnaire; SO: significant other.

**Table 2.**
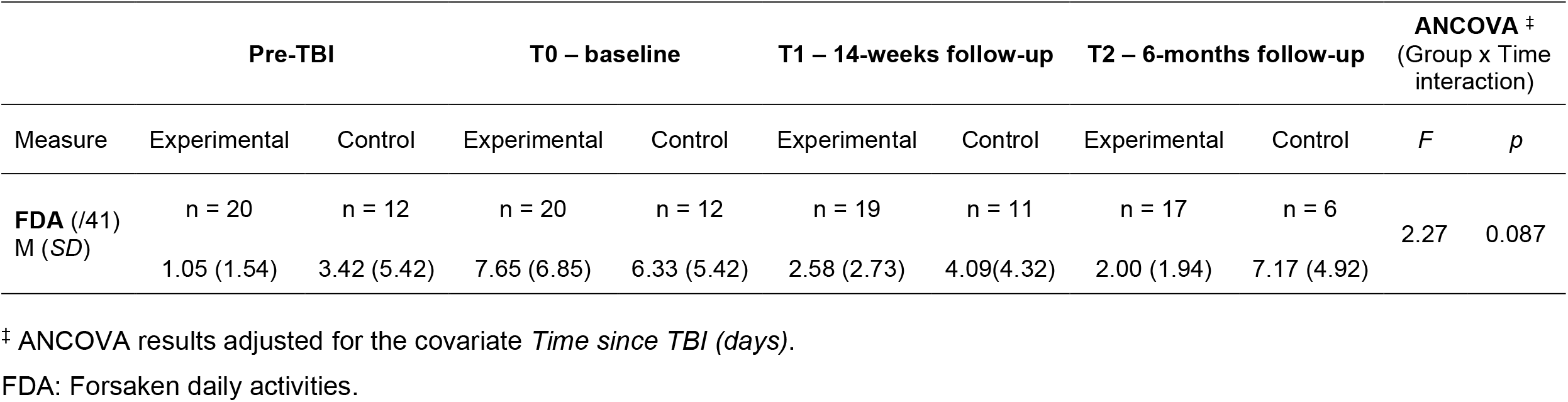
ANCOVA results for Forsaken Daily Activities (FDA)

### Primary outcome measures

There were no significant differences between experimental and active control groups before intervention, except for the SET-A variable ***Checking time*** (*t*(29) = 2.10, *p* = .04) where experimental participants showed higher scores than controls. Table 1 outlines means and standard deviations for the different measures at the three assessments times, and ANCOVA group-by-time interaction results after controlling for covariate *Time since TBI*.

There was a significant group-by-time interaction on ***Tackling the 6 subtasks*** from the SET-A with a moderate effect size. These significant differences were, however, not maintained at the 6-months follow-up, even though the score remained higher in the experimental group. Statistics for non-significant results not indicated in Table 1 are presented as Supplementary material. There were no significant main effects of time, or group on this measure. There was a significant main effect of time for ***Inter-task balance*** (*F*(2, 55) = 3.06, *p* = 0.05), but no significant main effect for group, nor a group-by-time interaction. A strong significant main effect of time was found for ***Checking time*** (*F*(2, 55) = 16.85, *p* < 0.000), as was a significant main effect for group (*F*(1,29) = 6.56, *p* = 0.016), but there was no significant group-by-time interaction. There was a significant group-by-time interaction on ***Avoiding rule-breaking*** with a moderate effect size. The pre-post-intervention difference for the experimental group was almost significant (*t* (28) = 1.94, *p* = 0.063; 95% CI [−2.23, 0.61]). These differences did not remain at the 6-months follow-up, even if ***Avoiding rule-breaking*** scores increased and remained higher in the experimental group. A significant main effect for group was also found for ***Efficient behavior*** (*F*(1, 29) = 4.24, *p* = 0.048), but no significant effect for time, nor group-by-time interaction were found. Supplementary bilateral t-test indicated that scores at T1 were significantly higher for the experimental group (*t* (29) = 2.35, *p* = 0.026; 95% CI [0.12, 1.75]). There was a significant main effect of time on ***SET-A Total Score*** (*F*(2, 42) = 7.23, p = 0.002), no main significant effect for group, and no group-by-time interaction. Supplementary t-test analysis indicated that scores at T1 were higher in the experimental group without quite attaining statistical significance (*t*(29) = 1.91, *p* = 0.06; 95% CI [−0.163, 4.618]). There were no significant effects for time or group, and no group-by-time interaction for ***Total number of points***.

There was no group-by-time interaction for ***Confirmed Correct Sorts*** total raw score of the ***Sorting Test***, nor a significant main effect for time, or group. Bilateral t-tests, however, indicated that scores at T1 were significantly larger for the experimental group than for controls (*t*(29) = 2.06, *p* = 0.048; 95% CI [0.12, 1.75]), but they were borderline significant six months later (*t*(22) = 1.96, *p* = 0.06; 95% CI [−1.09, 4.04]). For ***Free Sorting Description*** total raw score, ANCOVA showed no group-by-time interaction, no significant main effect for time, nor for group. Post-hoc bilateral t-tests show that higher scores in experimental group were borderline significant at T1 (*t*(29) = 1.96, *p* = 0.06; 95% CI [−.39, 17.85]), and significant at T2 follow-up (*t*(22) = 2.13, *p* = 0.04; 95% CI [0.22, 15.06]). A similar tendency was also found using a post-hoc ANOVA between the three assessment times (*F*(1, 21) = 4.04, *p* = 0.057). On the third *Sorting test* variable, ***Time-Per-Sort Ratio***, ANCOVA did not show a group-by-time interaction, there were no significant main effects for time or group. But post hoc bilateral t-test showed a significant difference at T1, the experimental group being faster to identify sorting categories (*t*(29) = - 2.65, *p* = 0.01; 95% CI [−24.38, −3.13]). Post-hoc ANOVA comparing T1-T0 also showed a significant difference favoring the experimental group (*F*(1, 21) = 5.02, *p* = 0.03), while ANOVA comparing the three assessment times showed a tendency in the direction of the hypothesis (*F*(1, 21) = 3.42, *p* = 0.078).

There was a strong significant main effect of time (*F*(2, 42) = 5.42, *p* = 0.008) on the ***Inhibition*** task of the ***Stroop*** four-color version, no significant main effect for group, and no group-by-time interaction. However, bilateral post hoc t-test showed significantly shorter time for the experimental group at T1 (*t*(28) = −2.45, *p* = 0.02; 95% CI [−56.66, −5.03]). There was a significant main effect for time on the ***Flexibility*** task (*F*(2, 42) = 3.19, *p* = 0.05), and for group (*F*(1, 21) = 4.42, *p* = 0.047) in favor of the experimental group, but no group-by-time interaction. Supplementary t-test showed a significant decrease in time to complete the ***Flexibility*** task at T1 (*t*(28) = −2.82, *p* = 0.009; 95% CI [−55.70, −8.79]). These results did not remain 6 months later.

### Generalization variables

Analyses showed a significant group-by-time interaction for participant-significant other difference scores on the ***Executive Cognition*** subscale of the ***DEX*** questionnaire. No significant main effect of time or group was found for this variable. Post-hoc t-tests showed borderline significant changes at T1 (*t*(22) = 1.92; *p* = 0.068, CI 95% [−0.64, 1.66]), and at T2 (*t*(23) = 1.78; *p* = 0.08, CI 95% [−0.94, 1.27]). As seen in Table 1, ***Behavioural-Emotional*** and ***Metacognitive*** subscales of the DEX did not show group-by-time interactions. A strong significant main effect for time was found for the ***Metacognitive*** subscale (*F*(2, 36) 17.30, *p* < 0.000), but no significant main effect was shown for group.

Participants identified ***Forsaken daily activities*** for four points in time, including at pre-injury. Results are presented in Table 2. A strong significant main effect for time was found (*F*(3, 76) = 11.68, *p* < 0.000). However, no group-by-time interaction and no significant group effect were identified. Post-hoc t-tests showed that before TBI, the control group reported more *Forsaken daily activities* than experimental participants although the difference did not quite reach significance (*t*(30) = −1.88, *p* = 0.07; 95% CI [−4.935, 0.202]). After TBI, at baseline T0, both groups reported significantly more *Forsaken daily activities*, thus reducing the differences between groups (*t*(30) = 0.57, *p* = 0.57; 95% CI [−3,430, 6.063]). After CEP intervention, at T1, even though the experimental group showed a reduction of *Forsaken daily activities* by more than half (−5.07) while control group reported a reduction of about 36% (−2.24), this difference was not statistically significant. Six months later, the differences between groups became highly significant (*t*(21) = −3.71, *p* = 0.001; 95% CI [−8.06, −2.27]), where the experimental group showed further reduction of *Forsaken daily activities* (−5.65 T2-T0), while they increased (+0.84 T2-T0) in control participants.

### Control measure

There were no significant changes on the ***Similarities*** subtest after CEP intervention.

## Discussion

This work aimed to contribute to filling a gap in clinical research on cognitive rehabilitation following TBI sustained by older adults. To our knowledge, this is the first study to address the effectiveness of a multimodal cognitive rehabilitation program developed for older adults with TBI, and its influence on enhancing executive functions. We developed a manualized rehabilitation program, the Cognitive Enrichment Program (CEP), from cognitive interventions having demonstrated positive effects in healthy older adults and younger adults with TBI. This paper reports on the effectiveness of the CEP program of executive functioning. Since aging and TBI increases variability, the CEP program consists in a system of organized cognitive rehabilitation strategies in order to address a diversity of rehabilitation needs. We compared an experimental TBI group with an active TBI group receiving usual care in the form of holistic rehabilitation without specific cognitive intervention. Our findings indicate that older adults with TBI are able to improve executive functioning as measured through ecologically sensitive executive function tests. Cognitive training also appeared to have facilitated resumption of everyday activities that had been abandoned since the TBI.

### Effectiveness of the CEP on executive functioning

Results show a significant improvement on two of measures of the Six Elements Task-Adapted: *Tackling the 6 subtasks* and *Avoiding rule-breaking*. These two measures are similar to the basic variables of the Behavioural Assessment of the Dysexecutive Syndrome (BADS) [37] scoring system. *Tackling the 6 subtasks* is a strategy allowing to rapidly earn more points on the test since approaching all sub-tasks in an equal fashion ensures solving the easier and higher.scoring items. This measure evaluates the participant’s ability to plan, elaborate and execute a main strategic solution in order to attain the main goal of the task. *Avoiding rule-breaking* demands keeping the rules active in working memory, self-monitoring, and updating the rule when necessary to inhibit an automatic response to switch to the immediate but wrong subtask. Improving both measures can reflect effectiveness of CEP interventions targeting EFs. The Planning Method (PM) [27] used taught participants to plan the steps and the resources (material, time, etc.), needed to solve a problem while being aware of contextual conditions of a task in order to be more efficient at the moment of actually conducting the task. The Problem.Solving Method (PSM) [27,28,30] used taught participants to consider as many possible solutions to a problem, select the best alternative, execute them, and assess their effect. This could have positively influenced the selection of the main strategy of approaching the 6 subtasks.

Goal Management Training (GMT) focused on self-monitoring, updating the goal and the chosen strategies, and adjusting it if necessary. Participant received a training to self-monitor progress and relevance of actual behavior compared to the goal and strategy while working on a task. In the SET-A, participants must self-monitor their behavior in order to frequently update their goal/rules and assess the effectiveness of their chosen strategy. In the particular case of *Avoiding rule-breaking*, GMT could have been useful to learn to update the rules at different moments of multitasking during the SET-A, by keeping endogenous attention on their own behavior, on the complex context of the task, and on keeping the rules in working memory. The strategies learned with GMT could also have trained participants to inhibit impulsive responses during the task.

*Checking the time* also seemed to be influenced by interventions, but intergroup comparison was affected by the fact that the experimental group was significantly better than controls before CEP, even though both groups showed problems in checking the time at baseline. Future research should consider this variable because it could be sensitive to EF interventions. Verifying the time at appropriate moments is one of the main dimensions of SET-A. Its prospective nature needs to keep endogenous divided attention on the task but also on the internal passage of time. Participant must elaborate a strategy to verify time at the most appropriate moments, keeping in working memory this rule and the strategy, update the rule and the strategy at the appropriate moments, to switch from the task to verify the time, and switch again to continue on the task. Because of its proven effectiveness in maintaining attention on the course of complex activities [18-20,25,26], it is probable that GMT could have positively influenced this specific strategy.

The experimental group showed a significantly higher score for *Efficient behavior* after CEP intervention on t-test analysis and ANCOVA showed a significant main effect of group. This variable is interesting because it represents a participants’ flexibility and adaptation ability to identify and use contextual information not directly mentioned in the instructions (organizing the task booklets, self-assessing one’s own speed on each subtask, etc.). The adaptive behaviors and participant’s engagement contribute to their main strategy. This is a measure of reduction of off-task behaviors that could negatively impact goal attainment, but also of on-task behaviors that contribute to achieving a goal. Although we were not able to show a significant group-by.time interaction for this variable, it is important to note that others did find significant changes on the task engagement of participants on an ecological task after GMT intervention [22,23].

The *Inter-task balance* measure did not appear to be very sensitive to EF interventions in both groups. The SET-A *Total Score* did generally improve throughout time and the experimental group showed a borderline significant improvement after the CEP intervention. On the other hand, the *Total number of points* measure did not show significant improvement after the CEP even if it should reflect general goal attainment, and as such, further research with this variable should be done to inform on its sensitivity.

ANCOVA failed to demonstrate significant effects on D-KEFS Sorting Test. However, supplementary t-test showed that the experimental group showed improvements on the three variables of the Sorting test after the CEP program, and this finding tended to remain six months later. It is interesting that at the 6-months follow-up we were able to detect slight further improvement on these three variables. Such measure should be included in future research since they assess categorization and cognitive flexibility. The PSM method used in the CEP emphasizes reasoning, logical thinking, and flexibility, and GMT targets flexibility and updating goals and strategies, which could influence efficacy on sorting tasks.

Results for Stroop measures failed to attain the significance criteria even if improvements were observed on both *Inhibition* and *Flexibility* variables after CEP interventions. Considering the limits of our study, we can’t be conclusive about these results. But these improvements and the fact that this task has been identified has a predictor of maintenance of independent living in elderly individuals [40] are encouraging for future research. Indeed, these authors demonstrated a direct relationship between executive performances and instrumental activities of daily living indicating predictive validity. Also, they proposed that cognitive training on EF areas may positively impact older adults’ performance on daily life activities.

### Generalization to real-life situations and daily activities

Since the CEP targets cognitive function, we hypothesized that changes could be seen on the *Executive Cognition subscale* of the DEX, but not on the other two subscales, which measure behavioural and emotional dimensions. Our findings support the hypothesis that CEP interventions have an impact on awareness of executive functioning by reducing the differences between the participant’s perception and their significant other’s perception of their executive functioning. Different strategies are integrated on the CEP program to approach different forms and degrees of unawareness. Also, feedback from the experimenter and from other participants, observation, vicariant learning, self-observation during homework, and other mechanisms implemented by the CEP could positively impact self-awareness, in particular in mild and moderate TBI. During intervention, special attention was put on constructive feedback about clinical improvement. This allowed the experimenter to highlight past unawareness while stressing improvements on self-awareness, and on cognitive progress.

The final goal of brain injury rehabilitation is the generalization of gains to daily life. In older TBI individuals this objective is more difficult to attain and measure since some older people are already reducing their activities in conjunction with their retirement. *Forsaken Daily Activities (FDA)* allowed us to assess activities normally abandoned before the TBI in order to compare them with those additionally abandoned after TBI and then following intervention. The number of *FDA* before TBI depends on highly interindividual variability and adaptive choices during aging. Research [40] has demonstrated the relationship between executive functioning and daily life activities in elders and the probable effect of cognitive training on improving daily life activities had been pointed out. Our findings are in this direction. Indeed, the experimental group resumed more than half their abandoned daily activities after intervention as compared to pre-intervention and, in a similar fashion as social roles in Spikman et al.’s study [28], they continue to improve six months later where the number of abandoned activities was not different from the number of activities already forsaken before the TBI, which is one of the main goals of rehabilitation. In fact, experimental participants had, in general, better outcomes than controls in the targeted executive measures used in this study, and even if all did not reach significance, they may have contributed to the resumption of daily life activities.

### Study limitations

Our study has some limitations which are already discussed in Cisneros et al. [29] (this issue). Attrition bias was one of the main factors of loss of statistical power and comparability of long.term effects, probably in detriment to the experimental group. Future clinical research must include larger and more stable samples by enrolling all patients at the beginning of their usual rehabilitation program to improve follow-up participation of older people with TBI. This will also reduce variability on time since injury. This was a pragmatic controlled trial which was controlled in an actual setting and as such, these parameters could not all be controlled.

## Conclusions

Our results show that older adults with TBI can improve their cognitive and daily life functioning after multimodal cognitive training, such as that as proposed by the CEP. This intervention program has demonstrated effectiveness for improving executive functioning, and also had an impact on daily functioning as measured by the resumption of daily life activities. As many researchers have pointed out, executive functions are good predictors of daily life functioning of older individuals, and this can be especially true following a TBI. Since older adults who sustain a TBI do show improvement in executive functioning after cognitive intervention, further clinical research focusing on rehabilitation strategies for this clinical population is warranted.

## Supporting information

ICMJE Form

## Data Availability

Data could be made available upon request.

## Acknowledgements

We wish to thank the TBI Programs of the Lucie-Bruneau and Le Bouclier Rehabilitation Centres, as well as the Trauma Program of the MUHC-MGH for their important role in participant recruitment. We also thank the individuals who participated in this research, sharing generously their time, efforts and energy. We would like to pay our gratitude and our respects to Dr. Donald T. Stuss, who, sadly, passed away in September 2019, and to Drs Gordon Winocur and Brian Levine for advice and encouragement at the early stages of this study, as well as to Drs Jacoba Spikman and Luciano Fasotti and their team in Holland for generously sharing and allowing translation of their Dysexecutive Training Protocol. We thank Dr. Jennifer Fleming for authorizing use and adaptation of her text *La conscience de soi*. We are grateful with the French artist Étienne Lécroart for allowing E.C. to use his ingenious cartoon *Perdre du temps* illustrating the negative impact of passivity. We thank Guylaine Bélizaire, Michel Ouellette, Christel Cornelis and Alexandra F. Girard who acted as research assistants for this project. We acknowledge the statistical work and advice of Miguel Chagnon. This project was funded by the Fonds de recherche du Québec – Santé (FRQS; research grant 22319 to M.M. and scholarship to E.C.), the Quebec Rehabilitation Research Network (research grant 09-10DS-06 to M.M.), the Lucie.Bruneau Rehabilitation Centre (research grant to E.C. and M.M., and clinical research hours to E.C.). The authors have no conflict of interest.

## Supplementary material

### Material and methods

#### Primary outcome measures

Crépeau, Belleville & Duchesne [32] developed a computerized version of the Six Elements Task [33], the Six Elements Task-Revised, allowing them to compare TBI patients to normal controls in seven aspects of the task: pre-planning time, number of tackled subtasks, maximum difference between parts of the same task, number of shifts between subtasks, rule-breaking, number of checks on timer, and ending time. The dictation task was substituted by a digit ordering task. They instructed the patient that the goal was to earn as many points as possible in 15 minutes. There were 30 items by subtask ordered with increasing difficulty but with decreasing scoring value. The patient stopped the time by themself. Our adaptation of Crépeau et al.’s [32] version to a paper-pencil format, named the ***Six Elements Task-Adapted (SET-A)***, included reducing the task completion time to 10 minutes, reducing the number of items to 25 per subtask aiming to be more suitable for older participants, adding counting of number of points earned in order to have a measure of general goal attainment. For the SET-A, five measures were thus used: ***Tackling the 6 subtasks, Inter-task balance*** (maximum item difference between parts of the same task), ***Avoiding rule-breaking, Checking time*** at appropriate moments (number of checks on timer and ending time), and ***Efficient behavior*** (adaptative/organized behavior during the test). Each measure represented a strategy contributing to attaining the main goal of earning as many points as possible. For each of these measures, we developed a scoring system to quantify the degree of attainment (0-3) of the optimal criteria (example: score of 2 if the person tackles 5 of 6 subtasks). We also calculated the ***Total score*** of the five measures of the SET-A (0-15), as well as the ***Total number of points*** earned (0-1950; 325 points for each subtask). All measures of SET-A showed significant positive correlations with SET-A total score (*Tackling the 6 subtasks r*(90) = .77, *p* < .001; *Inter.task balance r*(90) = .64, *p* < .001; *Checking time r*(90) = .59, *p* < .001; *Avoiding rule-breaking r*(90) = .29, *p* = .006; *Efficient behavior r*(90) = .59, *p* < .001; *Total number of points r*(90) = .63, *p* < .001). Cronbach alpha for the five measures was .45, which increased to .66 when excluding the *Avoiding rule-breaking* measure. We also validated the SET-A against the Wisconsin Card Sorting Test [34] measures in 92 healthy older adults, and showed moderate negative significant correlations between SET-A total Score and WCST Total correct responses (*r*(90) = - .33; *p* = .001), Number of trials (*r*(90) = .-27, *p* = .011), and Percent no perseverative responses (*r*(90) = −.22; *p* = .033).

In the ***Sorting Test*** [35], associated to problem-solving skills and concept formation, participants must sort six cards with printed words into two groups of three cards in as many ways as possible. The addition of correct sorts is the ***Confirmed Correct Sort Total (CCS)*** raw score. Participants then must verbalise the sorting criteria or category for each group, which represents the ***Free Sorting Description (FSD)*** total raw score, and ***Time-Per-Sort Ratio (TSR)*** is the mean (in seconds) invested to elaborate each sort.

The ***Stroop*** four-color version [36] contains four tasks (reading, colors, inhibition and flexibility), one hundred items by task, and four colors. The measures used (time, in seconds) were ***Inhibition*** of the Stroop effect by verbally indicating the color of the ink instead of reading the words, and ***Flexibility***, where participants indicated the color of the ink and read the words when they appeared in a box.

#### Generalization measures

The ***DEX*** [37,38] ***Executive cognition*** scale comprises items assessing temporal sequencing, planning, distractibility, and abstract thinking. The ***Behavioural-Emotional Self-Regulation*** scale is composed of items evaluating apathy, perseveration, lack of insight, confabulation, restlessness, variable motivation, knowing-doing dissociation, and lack of concern. The ***Metacognition*** scale is formed by items about aggression, euphoria, impulsivity, unconcern for social rules, inability to inhibit responses.

Since executive dysfunctions reduce independence in daily life activities, we considered that executive improvement could lead to increased resumption of daily activities that had been abandoned after the TBI. Self-reported ***Forsaken Daily Activities*** from the Client’s Intervention Priorities tool [39] were obtained for pre-injury and at each assessment time, where participants identified which of 41 daily activities they had abandoned.

## Results

### Primary outcome measures statistics for non-significant results

*SET-A - Tackling the 6 substasks*: pre-post-intervention difference not maintained at the 6-months follow-up (95% CI [−1.03, 0.90]); no significant main effect of time (*F*(2, 44) = 2.43, *p* = 0.10), or group (*F*(1, 22) = 0.21, *p* = 0.65).

*SET-A - Inter-task balance:* no significant main effect of group (*F*(1, 29) *= 0*.*87, p =* 0.36).

*Efficient behavior:* no significant effect of time (*F*(2, 52) = 2.17, *p* = 0.12).

*SET-A - Total score:* no significant main effect of group (*F*(1, 21) = 3.09, *p* = 0.09).

*SET-A - Total number of points:* no significant effect of time (*F*(2,40) = 2.12, *p* = 0.13), or group (*F*(1,20) = 0.61, *p* = 0.44).

*Sorting - Confirmed Correct Sorts* total raw score: no significant main effect for time (*F*(2, 57) = 0.43, *p* = 0.65), or group (*F*(1, 29) = 2.56, *p* = 0.12).

*Sorting - Free Sorting Description* total raw score: no significant main effect for time (*F* (2, 57) = 0.01, *p* = 0.99), nor for group (*F*(1, 20) 3.61, *p* = 0.06).

*Sorting - Time-per-Sort Ratio:* no significant main effect for time (*F*(2,57) = 0.52, *p* = 0.60), nor for group (*F*(1, 29) = 2.32, *p* = 0.14).

*Stroop - Inhibition*: no significant main effect for group, (*F*(1, 21) = 3.89, *p* = 0.60).

### Generalization measures statistics for non-significant results

*DEX - Executive Cognition* subscale: no significant main effect of time (*F*(2, 38) = 0.94, *p* = 0.40) or group (*F*(1, 24) = 2.10, *p* = 0.16).

*DEX - Metacognitive* subscale: no significant main effect of group (*F*(1, 24) = 3.53, *p* = 0.07). *Forsaken daily activities*: no significant group effect (*F*(1, 29) = 2.61, *p* = 0.11); no significant between-group T1-T0 reduction (*t*(28) = −1.18, *p* = 0.25; 95% CI [−4.142, 1.118]).

## TREND Statement Checklist

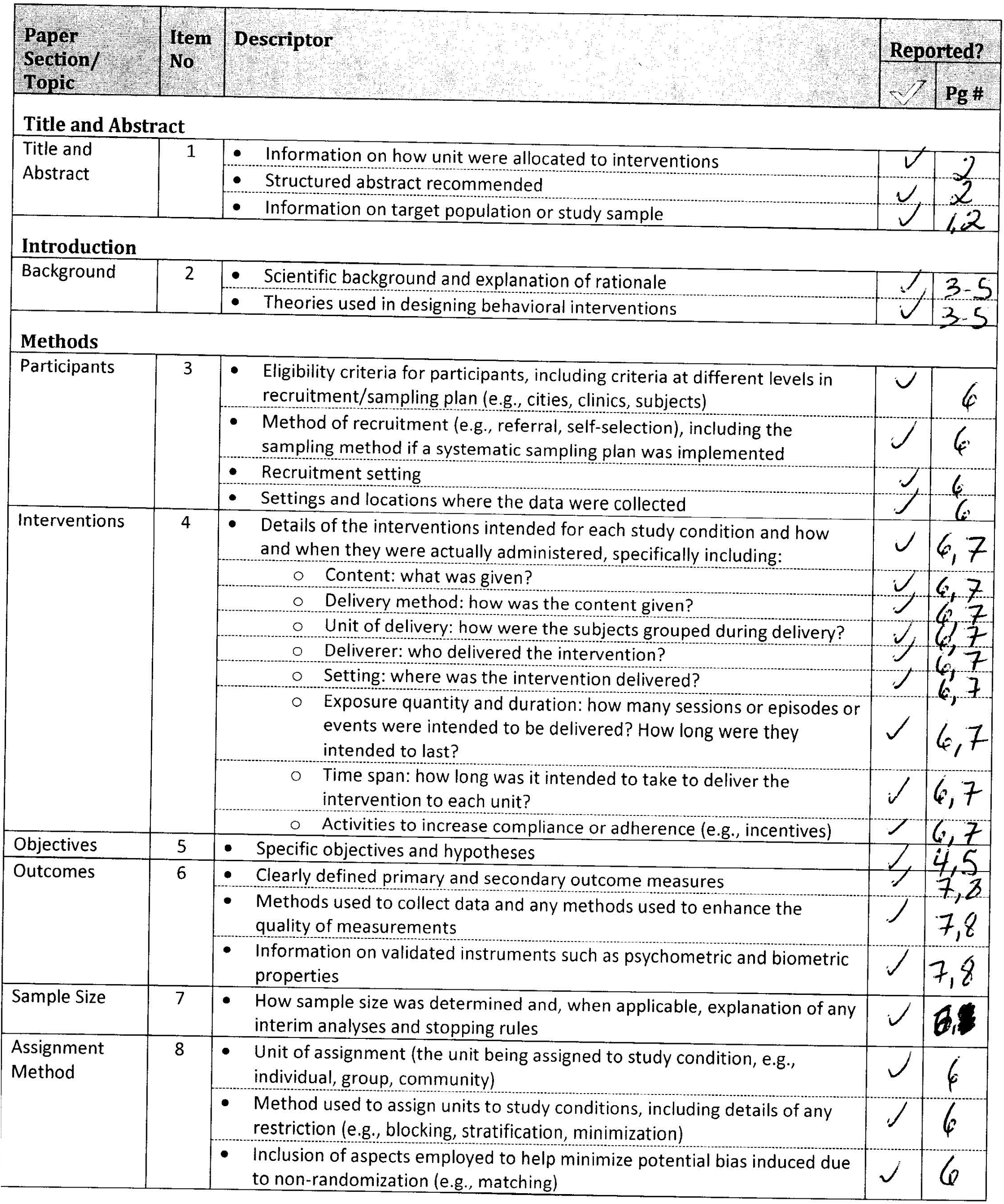
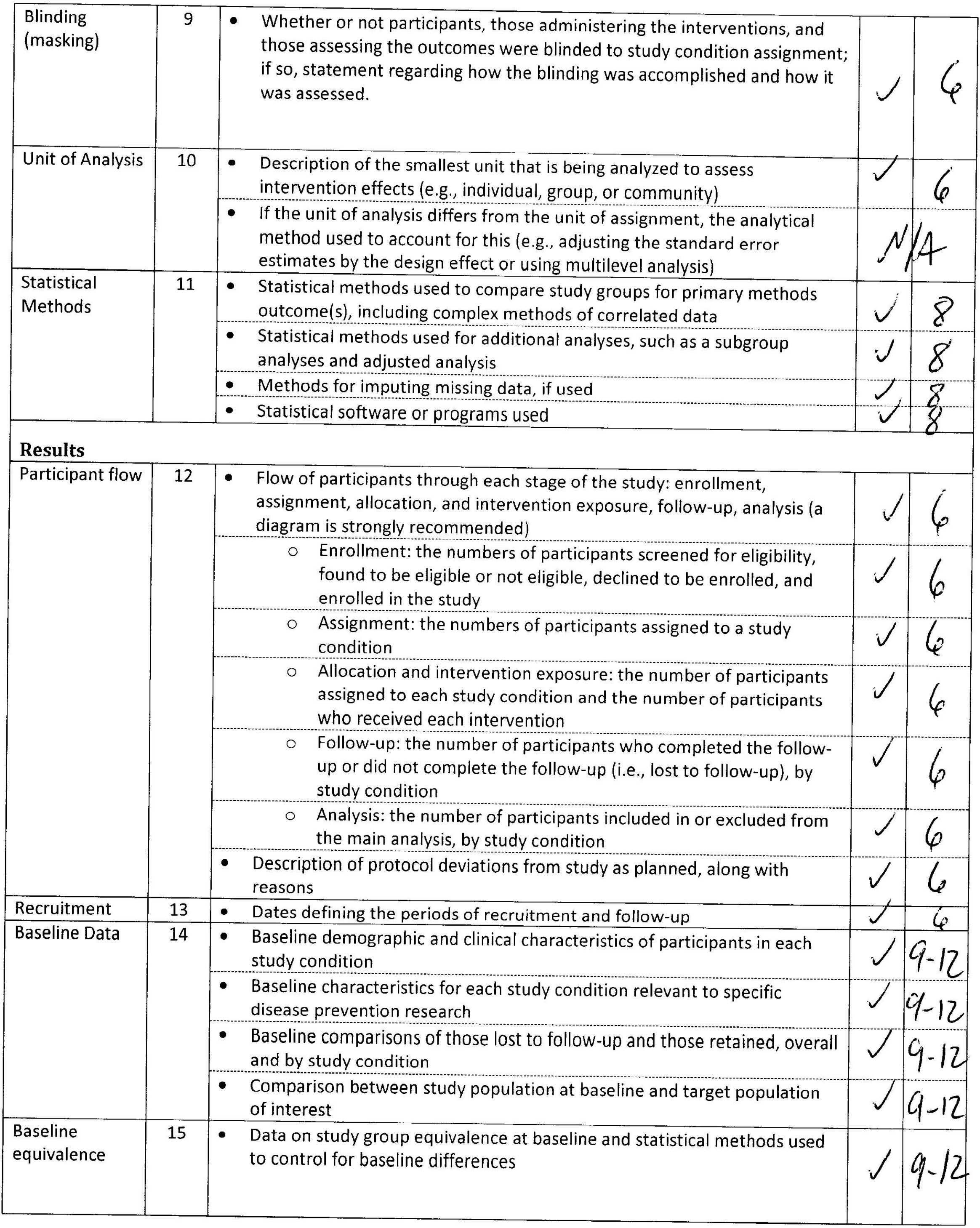
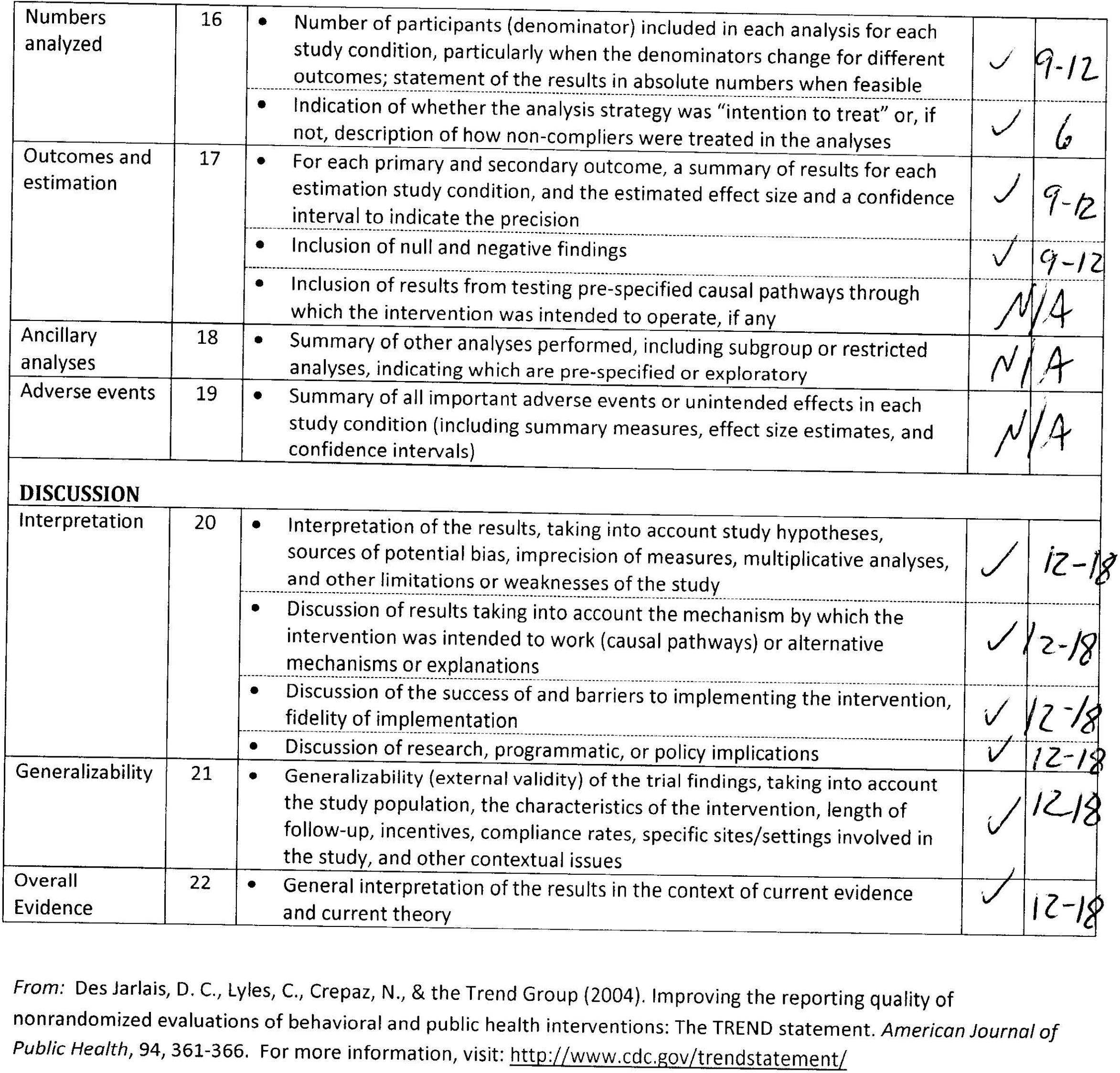

## Notes

### Competing Interest Statement

The authors have declared no competing interest.

### Clinical Trial

NCT04590911

### Funding Statement

Fonds de recherche du Quebec Sante (FRQS)
Quebec Rehabilitation Research Network
Lucie-Bruneau Rehabilitation Centre

### Author Declarations

CRIR and MUHC Ethical Review Boards

